# Serial Blood Microbiome Profiles in Kidney Transplant Recipients: Evidence of Circulating Gut Bacteria

**DOI:** 10.1101/2025.09.22.25330581

**Authors:** Alex DeVito, Ananda L. Kimm-Drapeau, William J. Higgins, Mia Montecillo, Carol Li, Isha Bal, Darshana M. Dadhania, Friederike Selbach, John R. Lee

## Abstract

Innovation in sequencing techniques has enabled the identification of microbiome in low biomass sites. In this study, we sought to investigate the utility of 16S rRNA deep sequencing of whole blood in kidney transplant recipients and to assess a link between the gut microbiota and the blood microbiota. We recruited 63 kidney transplant recipients who provided 163 whole blood specimens over the first 140 days after transplantation. We profiled the blood microbiome using 16S rRNA gene sequencing of the V4-V5 hypervariable region and additionally evaluated the gut microbiota in a subset of kidney transplant recipients who had gut bacteria detected in the blood. We generated a median of 19,959 sequences per blood specimen and found that most whole blood microbiome profiles consisted of mitochondrial DNA. While we did not identify classically pathogenic bacteria in the blood such as *Escherichia, Klebsiella*, and *Enterococcus*, we identified 25 gut bacterial taxa at very low levels in the blood of 22 kidney transplant recipients. For 9 of these kidney transplant recipients the same bacterial taxa detected in the blood were also identified in the gut microbiota. Our study is one of the first to show the detection of gut bacterial DNA in the blood of kidney transplant patients.

## Introduction

Newer sequencing techniques enable identification of the microbiome in low microbial biomass sites such as blood and urine. A cross-sectional study by Dominic Raj’s group described a diverse blood microbiome in patients with chronic kidney disease^1^. A pilot study with 6 kidney transplant recipients revealed a predominate abundance of Proteobacteria^2^. To date, however, no study has investigated the blood microbiome over the course of transplantation in kidney transplant recipients. The origins of bacterial taxa in the blood are thought to be, in part, from the gut. Indeed, several studies have shown that the source of bacteremia with pathogens such as *Enterococcus* can originate from the gut^3, 4^. Little, however, is known about whether gut commensal pathogens can be detected in the blood of patients.

The goal of our current study is to describe the utility of 16S rRNA deep sequencing of whole blood in kidney transplant recipients and to investigate a link between the gut microbiota and the blood microbiota in kidney transplant recipients.

## Methods

### Subject Recruitment

In this study, we recruited 63 kidney transplant recipients who received a kidney transplantation from October 2023 to June 2024 and who provided a total of 163 whole blood specimens over the first 140 days after transplantation. The Weill Cornell Institutional Review Board approved this protocol, and all subjects provided written informed consent. The study conforms to the principles of the Declaration of Helsinki. We considered this study as a pilot study and chose the above sample size. Within this cohort, 26 kidney transplant recipients provided two blood specimens and 37 kidney transplant recipients provided three blood specimens after transplantation. Whole blood specimens were stored directly at -80°C. Out of 22 kidney transplant recipients who had gut microbial taxa detected in the blood microbiota (gut microbial taxa were determined by the overlap of taxa among all of the fecal specimens from all of the patients with blood specimens), 19 patients had provided a total of 45 fecal specimens during the first 3 months after transplantation. Fecal specimens were obtained using the Fisherbrand stool commode kit and stored directly at -80°C.

### Blood Microbiome Sequencing and Bioinformatics

16S rRNA gene sequencing of the V4-V5 hypervariable region was performed on the blood specimens. Briefly, a whole blood aliquot was deposited into a Qiagen PowerBead glass 0.1 mm tube and was processed using a Promega Maxwell RSC PureFood GMO and Authentication Kit. The tube was vortexed, incubated at 60°C for 10 minutes, vortexed at high speed for 20 minutes, and centrifuged at 12,700 rpm for 10 minutes. The tube was then loaded into a Promega MaxPrep Liquid Handler tube rack to isolate DNA using a Promega MaxPrep Liquid Handler. 16S library generation was performed using the protocol from the Earth Microbiome Project. Pooled libraries were sequenced on an Illumina MiSeq Instrument (paired-end 250 base pair).

Raw reads were processed using Nextflow ^5^ nf-core^6^ ampliseq pipeline^7^, version 2.11.0. Taxonomic assignment was performed with DADA2, using the Silva reference database^8^.

### Gut Microbiome Sequencing and Bioinformatics

Whole genome sequencing was performed on the fecal specimens. Briefly, a single fecal specimen was deposited into a Qiagen PowerBead glass 0.1 mm tube and was processed using a Promega Maxwell RSC PureFood GMO and Authentication Kit. The tube was vortexed, incubated at 95°C for 5 minutes, vortexed at high speed for 20 minutes, and centrifuged at 12,700 rpm for 10 minutes. The tube was then loaded into a Promega MaxPrep Liquid Handler tube rack to isolate DNA using a Promega MaxPrep Liquid Handler. DNA was eluted in 100μl and transferred to a standard 96-well plate. Library generation was performed using the Illumina Nextera XT DNA Library Prep Kit. DNA sequencing libraries were washed using Beckman Coulter AMPure XP magnetic beads. Library quality and size verification was performed using the Revvity Labchip GX Touch HT instrument with DNA 1K Reagent Kit (CLS760673). Pooled libraries were sequenced on the Illumina NovaSeq X instrument at loading concentration of 0.13nM + 5% PhiX, paired-end 150bp.

Quality-based filtering and removal of host sequences and adapter and quality trimming was performed using kneadData (https://huttenhower.sph.harvard.edu/kneaddata/) version 0.10.0 using the hg37 genome. Taxonomic composition was calculated using MetaPhlAn4^9^. Gene families and pathways were renormalized to relative abundances after removing unmapped/ungrouped counts.

## Results

We recruited 63 kidney transplant recipients who provided 163 whole blood specimens over the first 140 days after transplantation. The mean age was 55±15 years old with 44% female kidney transplant recipients. Among the cohort, 32% were Caucasian, 25% were African American, 16% were Asian, and 27% were other or declined. 76% were non-Hispanic or Latino, 19% were Hispanic or Latino, and 5% declined.

We generated a total of 3,194,532 assigned sequences from the 163 whole blood specimens with a median of 19,959 sequences and interquartile range of 16,460 to 23,312 sequences. Importantly, we had two negative specimens for processing to assess for contamination through the sequencing processes and all specimens were analyzed on the same sequencing run to minimize batch variation. Among all the blood specimens, mitochondria DNA was the most classified taxa with a mean of greater than 99%. In subsequent analysis, we removed these mitochondria DNA taxa and we also importantly removed the 26 taxa that were identified in the 2 negative controls. Lastly, we removed any taxa that were not able to be identified at the genus level.

From the 163 blood specimens, 83 specimens from 51 kidney transplant recipients had detectable microbial 16S DNA classified with approximately 92 different taxa identified. Most taxa had an abundance of less than 1% within the blood specimens. Importantly, we did not identify many classically pathogenic bacteria in the blood such as *Escherichia, Klebsiella*, and *Enterococcus*. However, we did identify the following 25 gut bacterial taxa in 22 of the kidney transplant recipients’ blood microbiome profiles: *Acinetobacter, Akkermansia, Alistipes, Anaerococcus, Bacteroides, Blautia, Brevundimonas, Clostridium, Corynebacterium, Eubacterium, Faecalibacterium, Finegoldia, Lactobacillus, Lactococcus, Leptotrichia, Megasphaera, Prevotella, Pseudomonas, Romboutsia, Salmonella, Staphylococcus, Stenotrophomonas, Streptococcus, Turcibacter, and Veillonella*. Fig. 1 shows the relative abundance of these gut bacteria that were detected among all the blood specimens. We had 45 gut microbiota profiles available from 19 of the 22 kidney transplant recipients who had detectable gut bacterial taxa in their blood specimens. We found that the bacteria identified in the blood microbiome were identified in the gut microbiota in 9 of the 19 kidney transplant recipients (Fig. 2). Particularly, *Alistipes, Bacteroides, Blautia, Clostridium, Eubacterium, Romboutsia, Prevotella, Staphylococcus, Streptococcus*, and *Veillonella* were detected in both the blood and stool of the kidney transplant recipients (Fig. 2).

**Figure 1.**
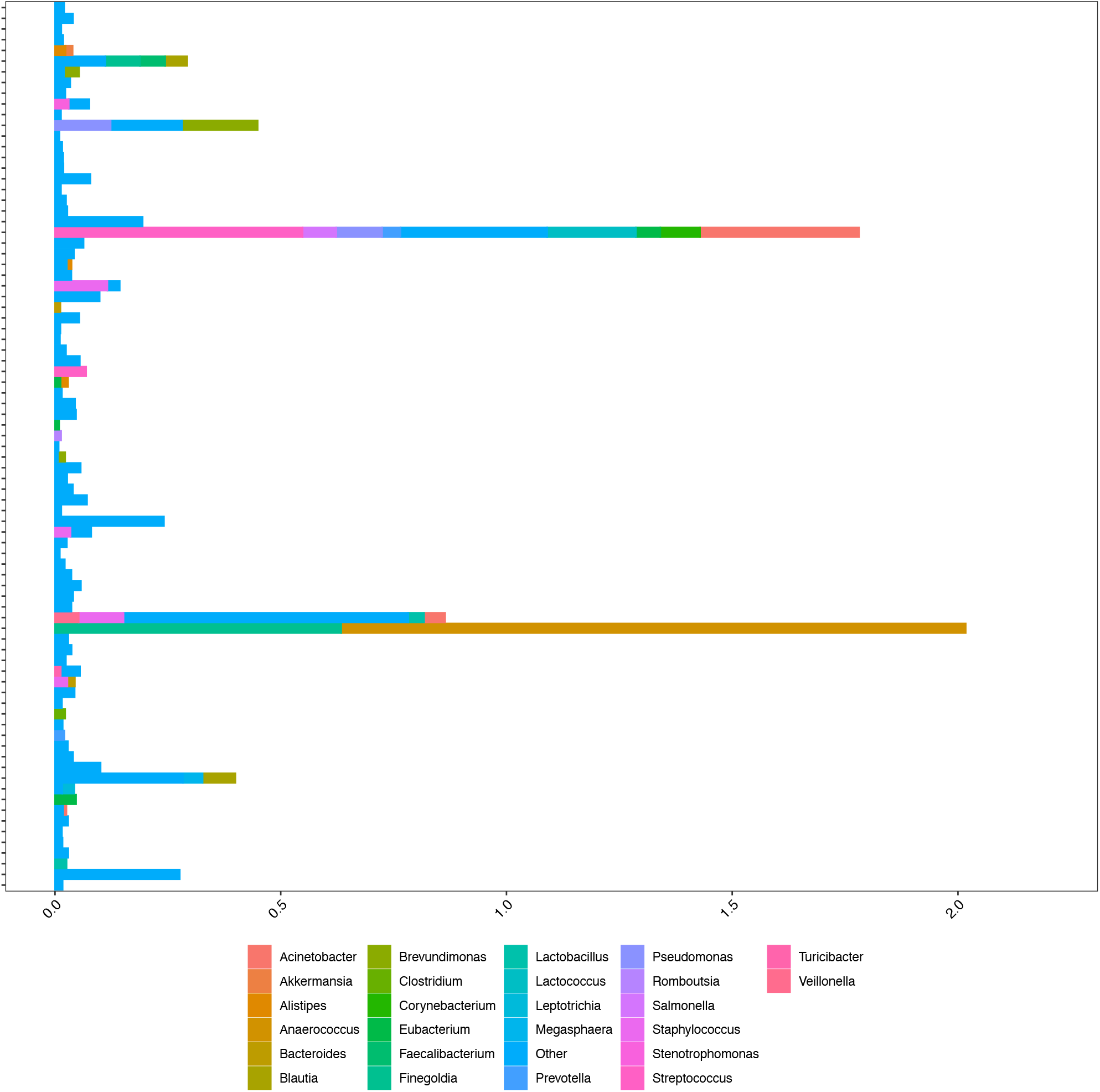
Gut Bacterial Taxa Detected in the Whole Blood of Kidney Transplant Recipients. The relative abundances of bacteria are represented as a percentage on the x-axis and each of the 81 16S blood profile from the kidney transplant recipients is shown on the y-axis. The bars represent the relative abundances of taxa whose identities are represented by the color in the legend.

**Figure 2.**
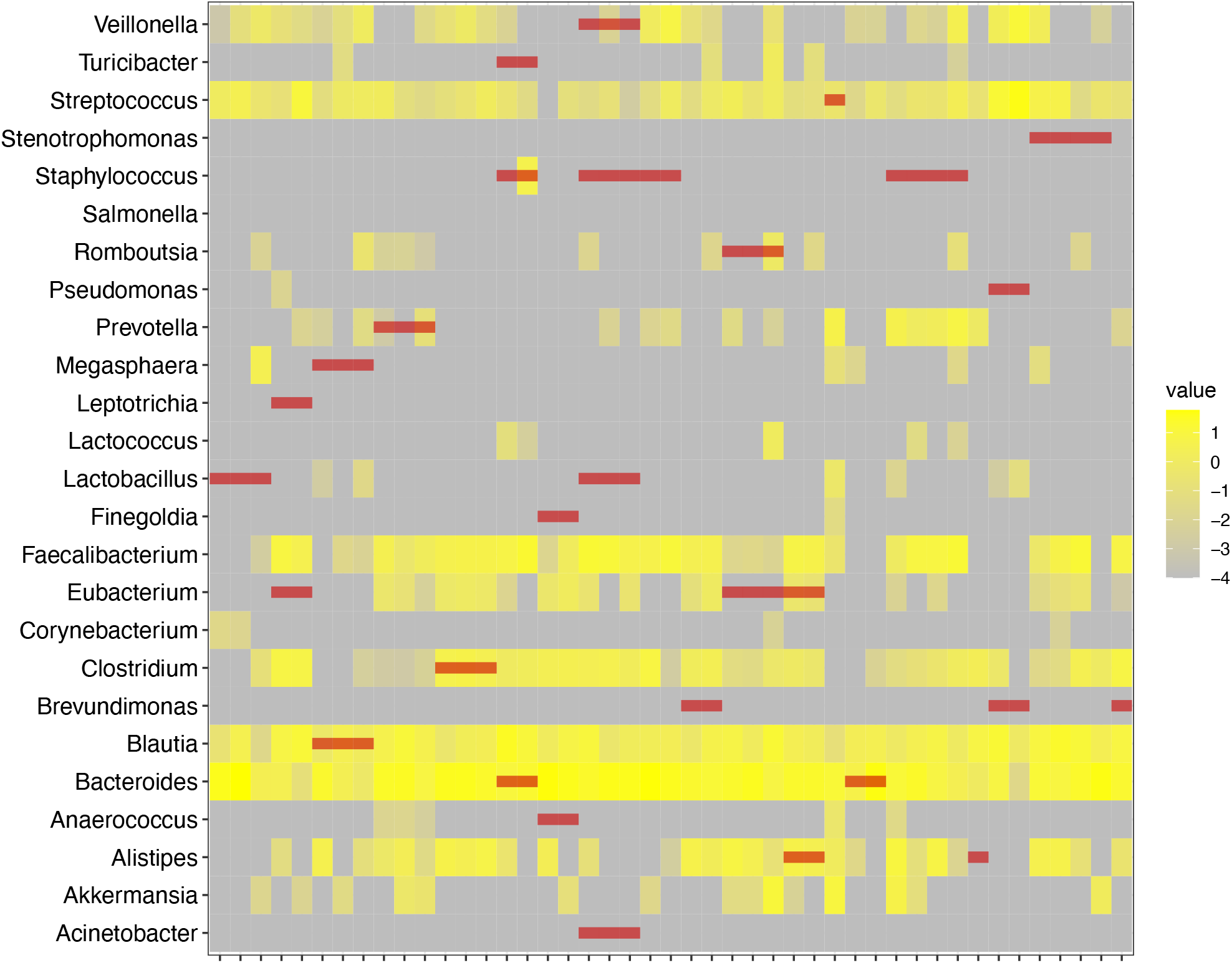
Overlap of Gut Bacterial Taxa in the Whole Blood and Gut Microbiota of Kidney Transplant Recipients. The heatmap represents the relative abundance of gut bacteria taxa in kidney transplant recipients as a percentage. On the x axis is the individual gut microbiota profiles from fecal specimens from kidney transplant recipients. On the y axis is individual gut bacteria taxa and the intensity is represented by varying shades of yellow on a log scale. The red lines represent bacteria taxa that were seen in the blood of kidney transplant recipients from the same subject.

## Conclusions

Our study provides one of the first descriptions of the blood microbiome over time in kidney transplant recipients. We first find minimal bacteria that is identified in the whole blood of kidney transplant recipients, while most sequencing reads stem from mitochondrial 16S DNA. Importantly, when compared to other blood microbiome studies, we performed this study on one sequencing run to minimize batch effects and potential bar hopping, and included negative controls. It is possible that sequencing a different compartment of the blood such as the buffy coat layer may have provided a more concentrated microbiome^10^. Nevertheless, our study provides novel information about sequencing circulating whole blood specimens in kidney transplant recipients and the finding that most DNA is attributable to mitochondrial DNA. To get more insight into the circulating 16S rRNA bacterial DNA, future studies will probably need to sequence at a deeper level.

Our study is also one of the first to show the detection of gut bacterial taxa in the blood of kidney transplant patients. Importantly, our study provided sequencing data of fecal specimens from the kidney transplant recipients to confirm that the identified gut bacterial taxa can be detected in the gut of the patients. While several studies have shown that gut pathogens like *Enterococcus* can translocate into the blood^3, 4^, our study shows that gut commensal bacteria can be detected in low levels of the blood. Indeed, a study by De Vlaminck et al. revealed that bacterial cell-free DNA can be detected in patients with inflammatory bowel disease^11^. Advanced methods like cell-free DNA profiling or deeper 16S sequencing may offer greater insight into gut bacteria via the blood, providing a more refined view of the gut microbiota in kidney transplant recipients.

## Data Availability

Sequencing data that support the findings of this study will be made available in the database of Genotypes and Phenotypes (dbGaP) phs004121.v1 upon publication.

## Authorship

AD: concept/design, collection of samples, data analysis/interpretation, critical revision of manuscript

ALK: concept/design, collection of samples, data analysis/interpretation, critical revision of manuscript

WJH: collection of samples, data analysis/interpretation, critical revision of manuscript

MM: collection of samples, data analysis/interpretation, critical revision of manuscript

CL: collection of samples, data analysis/interpretation, critical revision of manuscript

IB: collection of samples, data analysis/interpretation, critical revision of manuscript

DMD: concept/design, data analysis/interpretation, critical revision of manuscript

FS: data analysis/interpretation, critical revision of manuscript

JRL: concept/design, collection of samples, data analysis/interpretation, drafting article, critical revision of manuscript

## Acknowledgments

We thank Jorge Gandara, Alexander Grier, and the Weill Cornell Medicine Microbiome Core for their assistance in microbiome processing and analysis.

This manuscript was funded, in part, by National Institute of Health grant R21 AI 173849 (J.R.L.), National Institute of Health grant R01 DK 139249 (J.R.L.), and National Institutes of Health grant R01 AI 184528 (J.R.L.). F. S. is supported by research fellowship funding from the German Research Foundation (DFG, project number 539291308).

